# Neurological Disorders associated with COVID-19 Hospital Admissions : Experience of a Single Tertiary Healthcare Centre

**DOI:** 10.1101/2020.11.22.20235184

**Authors:** Permesh Singh Dhillon, Robert A. Dineen, Haley Morris, Radu Tanasescu, Esmaeil Nikfekr, Jonathan Evans, Cris S. Constantinescu, Akram A. Hosseini

**Affiliations:** Division of Clinical Neuroscience, University of Nottingham; Interventional Neuroradiology, Nottingham University Hospitals NHS Trust; NIHR Nottingham Biomedical Research Centre; Department of Neurology, Queen’s Medical Centre, Nottingham University Hospitals NHS Trust, United Kingdom

**Keywords:** COVID-19, encephalopathy, neurology, delirium, stroke

## Abstract

**Background:** Early reports have detailed a range of neurological symptoms in patients with the SARS-CoV-2 infection. However, there is a lack of detailed description and incidence of the neurological disorders amongst hospitalized COVID-19 patients. We describe a range of neurological disorders (other than non-specific neurological symptoms), including their clinical, radiological and laboratory findings, encountered in our cohort of COVID-19 patients admitted to a large tertiary institution.

**Methods:** We reviewed our prospectively collated database of all adult Neurology referrals, Neurology and Stroke admissions and Neurological multi-disciplinary team meetings for all hospitalized patients with suspected or proven COVID-19 from 17 March 2020 to 31 August 2020.

**Results:** Twenty-nine of 1243 COVID-19 inpatients (2.3%) presented with COVID-19-related neurological disorders. The mean age was 68.9 +/- 13.5(SD) years, age range of 34-97 years, and there were 16 males. 22 patients had confirmed, 5 were probable and 2 had suspected COVID-19 infection according to the WHO case classification. Eight patients (27%) required critical care admission. Neurological symptoms at presentation included acute confusion and delirium, seizures, and new focal neurological deficits. Based on the pre-defined neurological phenotype, COVID-19 patients were grouped into four main categories. 16 patients had cerebrovascular events (13 with acute ischaemic stroke and 3 had haemorrhagic features), 7 patients were found to have inflammatory, non-inflammatory and autoimmune encephalopathy (including 2 with known Multiple Sclerosis), whilst disorders of movement and peripheral nervous system were diagnosed in 3 patients each.

**Conclusion:** Although the exact prevalence and aetiology remain unclear, new onset of neurological disorders, in addition to anosmia, is non-sporadic during the acute COVID-19-infection. Longitudinal follow-up of these patients is required to determine the clinical and functional outcome, treatment response and long-term effects of the SARS-CoV-2 infection.

## Introduction

The coronavirus disease 2019 (COVID-19), a manifestation of the severe acute respiratory syndrome coronavirus-2 (SARS-CoV-2), was declared a pandemic by the World Health Organisation (WHO) on 11 March 2020 (1, 2). At present, the COVID-19 incidence in the United Kingdom (UK) is one of highest in the world with 705,428 cases and 43,579 deaths, accurate as of 17 October 2020 (3). Early reports from Wuhan, China detailed a range of neurological symptoms seen in patients with the SARS-CoV-2 infection (4). Recent isolated case reports have also described some of these manifestations, which include acute cerebrovascular disorders [CVD] (5-7), encephalopathy or encephalitis, acute demyelinating encephalomyelitis (ADEM), as well as peripheral neurological associations such as Guillain-Barre syndrome (GBS) (8, 9). Some of the proposed mechanisms underlying the increased prevalence of neurological disorders in COVID-19 include widespread systemic inflammatory and cytokine responses, diffuse intravascular coagulation and/or critical illness-related coagulopathy, direct neuronal injury, immune-mediated disorders and haemodynamic alterations (8-14).

Many reports have detailed a range of presenting neurological symptoms in patients with the SARS-CoV-2 infection, including headache, delirium, seizures and altered mental status. However, there is often a lack of detailed description and incidence of the neurological disorders amongst hospitalized patients with COVID-19. Herein, we solely include neurological disorders, instead of non-specific neurological symptoms such as headache, dizziness and anosmia. We describe a range of neurological disorders causing neurological deficits, including their clinical, radiological and laboratory findings, encountered in our cohort of patients with COVID-19 admitted to a large tertiary institution.

## Methods

This study was registered with and approved by the East Midlands-Derby Research Ethics Committee (Ref:18/EM/0292, Major amendments) and individual patient consent was waived (15). We reviewed our prospectively collated database of all inpatient Neurology referrals, Neurology and Stroke admissions and Neurological multi-disciplinary team (MDT) meetings for all hospitalized patients with suspected or proven COVID-19 from 17 March 2020 (when national lockdown was declared in the UK) to 31 August 2020, at our institution. Each case, including the clinical, laboratory, and imaging findings, was discussed and a consensus of the underlying COVID-19 associated neurological syndrome was reached amongst the Neurology or Stroke physicians. Cases without definite neurological deficits, symptoms or signs, no clinical/radiological suspicion of COVID-19, or other more likely alternate diagnoses were excluded from our study cohort.

Patients presenting with symptoms and/or signs indicative of COVID-19 and the associated positive real-time reverse transcriptase polymerase chain reaction (RT-PCR) status from the naso-pharyngeal swab test, were classified according to the WHO COVID-19 case definition (16), into confirmed, probable and suspected cases. Further case definitions for the association of COVID-19 with neurological disease were defined based our local MDT consensus and adapted from Ellul et. al’s compilation panel of published information (14). These included cerebrovascular disease, encephalitis, myelitis or meningitis, and acute disseminated encephalomyelitis or other acute neuropathies associated with the SARS-CoV-2 infection. Some findings of four patients from our cohort were described in a recent correspondence by the respective clinicians/authors : (Patients 21 and 22) by Hosseini et al. (17) and (Patients 18 and 28) by Dhillon et al. (18).

## Results

Our tertiary institution holds a capacity of approximately 1700 hospitals beds and provides services to over 2.5 million residents. During the study period, our institution reported 1243 COVID-19 admissions, of which 29 patients (2.3%) with neurological disorders associated with COVID-19 were identified in our study. The 29 patients included had a mean age of 68.9 +/- 13.5(SD) years, age range of 34-97 years, and there were 16 males. There were 27 Caucasian patients (92%), and only 2 were from the Black, Asian or Minor Ethnicity groups (BAME ; 1 Black and 1 Asian). According to the WHO COVID-19 case classification, 22 patients were deemed to have confirmed COVID-19, 5 were probable and 2 had suspected COVID-19 infection. Eight patients (27%) required critical care admission, 6 of whom needed invasive ventilation. There was an array of neurological symptoms at presentation, namely, reduced consciousness, acute confusion, behavioural change and seizures, acute motor or sensory neurological deficits, and acute onset of movement disorders. The onset of neurological symptoms was between 9 days before to 15 days after the diagnosis or symptoms onset of COVID-19. Based on the pre-defined neurological phenotype, COVID-19 patients were grouped into four main categories : 16 patients diagnosed with a Cerebrovascular event (Acute ischaemic and Haemorrhagic), 7 patients with Inflammatory, non-inflammatory and autoimmune encephalopathy (including one case of transverse myelitis and two cases with known Multiple Sclerosis), 3 patients with Movement disorders, and 3 patients Peripheral nervous system syndromes.

### Cerebrovascular Event

#### Acute Ischaemic Stroke

(Patient 1, 2, 3, 4, 6, 10, 11, 12, 15, 16, 25, 29, 30; Table 1)

**Table 1.** Sixteen patients with cerebrovascular events (13 Ischaemic stroke and 3 Haemorrhagic).

| Patient | 1 | 2 | 3 | 4 | 6 (Figure 1) |
| --- | --- | --- | --- | --- | --- |
| Age, M/F, ethnicity, COVID-19 diagnosis | 90s, F, White, Definite | 90s,F, White, Definite | 60s,F, White, Definite | 80s, M, White, Probable | 70s, M, White, Definite |
| Stroke type, thrombolysis/thrombectomy | Left ICA/MCA infarct. Thrombolysis | Left partial anterior circulation stroke (PACS) | Left ICA/MCA infarct ; Mechanical thrombectomy | Right MCA infarct. Thrombolysis | Right MCA infarct |
| Blood results at admission: | Haemoglobin 136 g/l<br>lymphocyte count 1.39<br>neutrophil count 3.41<br>platelet count 207<br>CRP 6 mg/L<br>D-dimer; NR<br>Ferritin; NR<br>Creatinine 66 umol/L<br>PT 12.6 seconds | Haemoglobin 125 g/l<br>**lymphocyte count 0.98**<br>neutrophil count 10.98<br>platelet count 421<br>**CRP 276 mg/L**<br>**ESR mm/hr: 116**<br>D-dimer; NR<br>Ferritin; NR<br>Creatinine 59 umol/L<br>PT 11.1 seconds | Haemoglobin 131 g/l<br>lymphocyte count 2.91<br>neutrophil count 9.74<br>platelet count 400<br>**CRP 62 mg/L**<br>**D-dimer ug/l; 3144**<br>Ferritin; NR<br>Creatinine 64 umol/L<br>PT 11.6 seconds | Haemoglobin 140 g/l<br>**lymphocyte count 0.92**<br>neutrophil count 5.53<br>platelet count 231<br>**CRP 23 mg/L**<br>D-dimer; NR<br>Ferritin; NR<br>Creatinine 89 umol/L<br>PT 11.9 seconds | Haemoglobin 106 g/l<br>**lymphocyte count 0.15**<br>neutrophil count 2.81<br>**platelet count 38**<br>**CRP 298 mg/L**<br>D-dimer; NR<br>Ferritin; NR<br>Creatinine 119 umol/L<br>PT 11.9 seconds |
| Brain Imaging | CT: Acute thrombus within the left ICA and MCA consistent with an acute infarct | CT: No acute abnormality. Severe small vessel disease. | CT: Left MCA territory infarct, with associated mass effect. | CT: Right MCA territory infarct. | CT: Massive acute right MCA territory infarct with mass effect and shift of midline structures |
| Outcome status | Died | Full recovery | Continued recovery in rehabilitation ; mRS 5 | Full recovery | Died |

| 10 | 11 | 12 | 15 | 16 | 25 |
| --- | --- | --- | --- | --- | --- |
| 80s, F, White, Definite | 60s, F, White, Probable | 70s, M, White, Suspected | 60s, M, White, Definite | 80s, M, White, Definite | 70s, F, White, Definite |
| Right PCA Infarct | Left ACA infarct | Right MCA infarct | Right MCA infarct | Right MCA infarct | Left MCA infarct ; Mechanical Thrombectomy |
| Haemoglobin 113 g/l<br>**lymphocyte count 0.80**<br>neutrophil count 6.82<br>platelet count 296<br>**CRP 30** mg/L<br>D-dimer; NR<br>Ferritin 126 ug/l<br>Creatinine 62 umol/L<br>PT; NR | Haemoglobin 108 g/l<br>**lymphocyte count 0.76**<br>neutrophil count 11.27<br>platelet count 196<br>**CRP 74** mg/L<br>D-dimer; NR<br>Ferritin; NR<br>Creatinine 34 umol/L<br>PT 12.1 seconds | Haemoglobin 145 g/l<br>**lymphocyte count 0.49**<br>neutrophil count 8.14<br>platelet count 332<br>**CRP 132** mg/L<br>D-dimer; NR<br>Ferritin 38 ug/l<br>Creatinine 81 umol/L<br>PT 13 seconds | Haemoglobin 117 g/l<br>**lymphocyte count 0.87**<br>neutrophil count 10.72<br>platelet count 352<br>**CRP 296** mg/L<br>D-dimer; NR<br>Ferritin; NR<br>Creatinine 84 umol/L<br>PT 10.9 seconds | Haemoglobin 136 g/l<br>**lymphocyte count 0.82**<br>neutrophil count 7.87<br>platelet count 335<br>**CRP 345** mg/L<br>D-dimer; NR<br>Ferritin; NR<br>**Creatinine 118** umol/L<br>PT 14.4 seconds | Haemoglobin 105 g/l<br>lymphocyte count 1.19<br>neutrophil count 8.49<br>platelet count 362<br>CRP 8 mg/L<br>D-dimer; NR<br>Ferritin; NR<br>**Creatinine 106** umol/L<br>PT 12.6 seconds |
| MRI: Acute right PCA infarct | CT: Acute left ACA territory infarct | CT: Small cortical infarcts involving the right pre-central gyrus. | CT: Large acute right MCA territory infarct with mass effect | CT; Acute thrombus within the terminal segment ICA, M1/M2 segments of the right MCA and the proximal right A1 ACA segment | CT : Left MCA thrombus and infarct |
| Continued recovery in rehabilitation ; mRS 3 | Full recovery | Continued recovery in rehabilitation ; mRS 3 | Died | Died | Full recovery |

| 28 (Figure 1) | 18 (Figure 1) | 24 | 29 | 30 |
| --- | --- | --- | --- | --- |
| 60s, M, White, Definite | 40s, M, White, Definite | 50s, F, BAME, Suspected | 30s, F, White, Definite | 60s, M, Definite |
| Isolated intraventricular haemorrhage | 1. Isolated intraventricular haemorrhage<br>2. Unilateral hearing loss (new) | Cerebral microbleeds | Multifocal infarct | Multifocal infarcts |
| Haemoglobin 127 g/l<br>**lymphocyte count 0.17**<br>neutrophil count 3.99<br>platelet count 228<br>**CRP 158** mg/L<br>**D-dimer 8645** ug/l<br>**Ferritin 758** ug/l<br>**Creatinine 128** umol/L<br>PT 10.8 seconds | Haemoglobin 141 g/l<br>**lymphocyte count 0.52**<br>neutrophil count 3.51<br>platelet count 170<br>**CRP 171** mg/L<br>D-dimer; NR<br>**Ferritin 2132** ug/l<br>Creatinine 91 umol/L<br>PT 11.4 seconds | Haemoglobin 147 g/l<br>**lymphocyte count 0.86**<br>neutrophil count 6.02<br>platelet count 202<br>CRP <5 mg/L<br>D-dimer; NR<br>Ferritin; NR<br>Creatinine 95 umol/L<br>PT 12.9 seconds | Haemoglobin 91 g/l<br>**lymphocyte count 0.94**<br>neutrophil count 5.87<br>platelet count 455<br>**CRP 263** mg/L<br>**D-dimer 6358** ug/l<br>**Ferritin 2986** ug/l<br>**Creatinine 286** umol/L<br>PT 11.4 seconds | Haemoglobin 132 g/l<br>**lymphocyte count 0.66**<br>neutrophil count 9.35<br>platelet count 412<br>**CRP 342** mg/L<br>D-dimer; NR<br>**Ferritin 3712** ug/l<br>Creatinine 75 umol/L<br>**PT 13.1** seconds |
| CT; Small amount of intraventricular haemorrhage in the left occipital horn of the lateral ventricle. | CT and MRI: Small volume intraventricular haemorrhage layering in the occipital horns bilaterally. | MRI; Micro-haemorrhagic changes in both cerebral hemispheres | CT and MRI: Multiple bilateral white matter and left cerebellar tiny foci of infarction | CT; Multifocal supra- and infra-tentorial infarcts. |
| Full recovery | Recovery with slight disability; mRS 1 | Full recovery | Died | Died |

Footnote : M = male, F = female, ICA = internal carotid artery, ACA = anterior cerebral artery, MCA = middle cerebral artery, PCA = posterior cerebral artery, NR = no result, CRP = C-Reactive protein, ESR = Erythrocyte sedimentation rate, mRS = modified Rankin Score, PT = Prothrombin time, CT = computed tomography. MRI = magnetic resonance imaging, Lymphocyte, neutrophil and platelet count; numbers x 10 E9/L.

Thirteen of the 28 patients (46%) with an age range of 34-97 years, and 7 females, were diagnosed with acute ischaemic stroke, 5 of which were large vessel occlusions involving the middle cerebral artery (MCA). Only one patient presented with a posterior circulation stroke whilst two had multifocal infarcts. All but one patient had at least one known cardiovascular risk factor. Two patients were admitted in the intensive care unit for management of a malignant MCA syndrome. All patients underwent a computed tomography (CT) and/or an MRI Head at presentation (representative examples in Figure 1). Only two patients were given intravenous (IV) thrombolysis and two underwent mechanical thrombectomy (MT). Five patients died within days of their diagnosis due to a combination of the underlying stroke and/or COVID-19 pneumonia.

**Figure 1:**
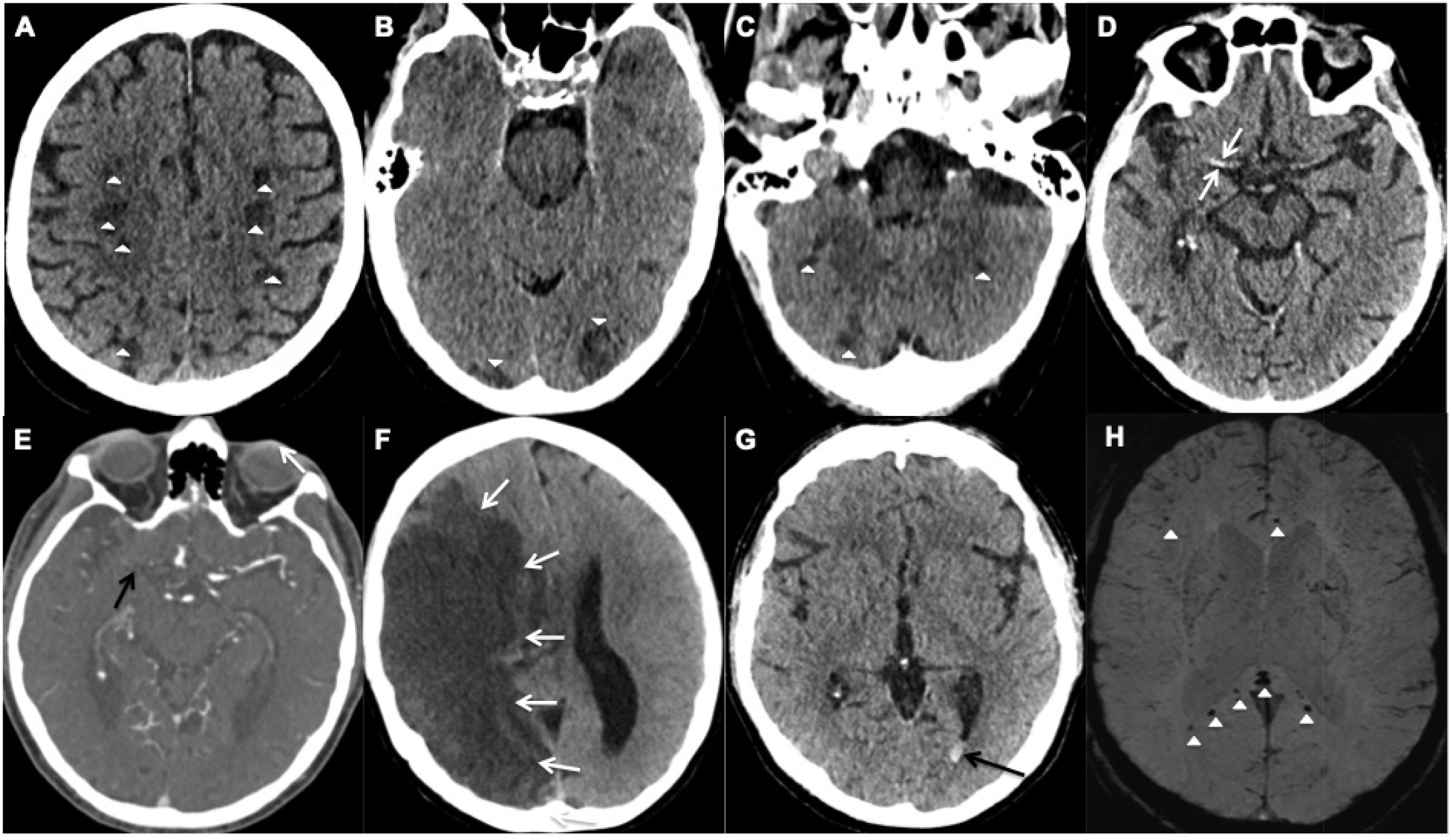
Representative examples of cerebrovascular events; (A-F) acute ischaemic stroke and (G-H) haemorrhagic events. (A-C) Axial unenhanced CT images of Patient 28 demonstrate multifocal infarcts (white arrowheads) bilaterally along the (A) centrum semiovale, (B) Occipital lobes and (C) Cerebellar hemispheres. (D-E) Axial CT images of Patient 25 demonstrates the (D) hyperdense right middle cerebral artery (MCA) sign (white arrows) and (E) corresponding filling defect on the CT angiogram confirming an occlusive thrombus (black arrow). (F) Axial unenhanced CT image of Patient 6 shows a large right MCA territory infarct (white arrows) with mass effect in keeping with an MCA malignant syndrome. (G-H) Axial images of Patient 18. (G) unenhanced CT demonstrates hyperdense layering in the occipital horn of the left lateral ventricle (black arrow) in keeping with isolated intraventricular haemorrhage (IVH), (H) Susceptibility weighted imaging confirms the IVH bleed within the left occipital horn (white arrow) and shows microbleeds at the splenium and genu of the corpus callosum, and subcortical white matter (white arrowheads).

#### Haemorrhagic

(Patient 18, 24, 28; Table 1)

Two COVID-19 male patients, Patients 18 and 28, (aged in their 40s and 60s respectively) had isolated intraventricular haemorrhage (IIVH) demonstrated on the CT Head, performed due to reduced conscious levels despite a sedation hold, during their prolonged critical care admission. MRI Head findings of the 48-year-old patient showed cerebral microbleeds (CMB) in the splenium of the corpus callosum and subcortical white matter (Figure 1). Both patients were placed on continuous veno-venous haemofiltration (CVVH) but neither required extracorporeal membrane oxygenation (ECMO). The platelet count level, prothrombin and activated partial thromboplastin times were within the normal referenced ranges.

The third patient in her 50s (Patient 24) presented with recurrent seizures. The MRI Head also revealed CMB, without any other diagnostic features of cerebral amyloid angiopathy. No critical care admission was required and the prothrombin time was normal. Two patients (Patient 24 and 28) made an uneventful recovery. However, patient 18 reported unilateral hearing impairment following hospital discharge.

### Inflammatory, Non-inflammatory and Autoimmune Encephalopathy

Limbic encephalitis (Patient 21); Inflammatory encephalopathy (Patient 22); ADEM (Patient 27); Transverse myelitis (Patient 5); Non-inflammatory encephalopathy (Patient 9); (Table 2)

**Table 2.** Seven patients with inflammatory, non-inflammatory or autoimmune encephalopathy, including two patients with known multiple sclerosis (MS), and one patient with transverse myelitis.

| Patient | 27 (Figure 2) | 21 (Figure 2) | 22 | 5 | 9 | 8 | 14 |
| --- | --- | --- | --- | --- | --- | --- | --- |
| Age, M/F, ethnicity, COVID-19 diagnosis | 60s, F, White, Definite | 70s, F, White, Definite | 40s, M, BAME, Definite | 60s, M, White, Probable | 50s, F, White, Definite | 60s, M, White, Definite | 50s, M, White, Definite |
| Final neurological diagnosis (impression) | Acute demyelinating encephalomyelitis (ADEM) | Limbic encephalitis associated with SARS-CoV2 | Inflammatory encephalopathy associated with SARS-CoV2 | Transverse myelitis | Non-inflammatory encephalopathy associated with SARS-CoV2 | Relapse in an advanced Secondary Progressive MS | Relapse in a known Multiple Sclerosis |
| Key neurological signs | Delirium, limb weakness, ataxia, visual hallucination, | Delirium, New onset generalised seizure/status epilepticus, dysphagia, cognitive impairment, amnesia | Delirium, New onset generalised seizures, disinhibition and cognitive impairment | Quadriparesis and sensory loss at cervical level | Headache, delirium, reduced conscious level, confusion and behavioural change | Delirium, reduced consciousness | Worsening limb weakness and dysarthria |
| Blood results at admission: | Haemoglobin 124 g/l<br>**lymphocyte count 0.55**<br>neutrophil count 9.11<br>platelet count 336<br>**CRP 262 mg/L**<br>ESR 10 mm/hr<br>D-dimer; NR<br>Ferritin; NR<br>Creatinine 94 umol/L<br>PT 10 seconds | Haemoglobin 132 g/l<br>**lymphocyte count 0.39**<br>neutrophil count 5.83<br>platelet count 313<br>**CRP 31 mg/L**<br>**D-dimer 3748 ug/l**<br>Ferritin; NR<br>Creatinine 65 umol/L<br>PT 13.7 seconds | Haemoglobin 147 g/l<br>**lymphocyte count 0.89**<br>neutrophil count 3.6<br>platelet count 133<br>**CRP 149 mg/L**<br>**ESR 90 mm/hr**<br>**D-dimer 1659 ug/l**<br>**Ferritin; 1328 ug/l**<br>Creatinine 88 umol/L<br>PT 11.9 seconds | Haemoglobin 120 g/l<br>lymphocyte count 1.1<br>neutrophil count 8.9<br>platelet count 407<br>**CRP 24 mg/L**<br>D-dimer; NR<br>Ferritin; NR<br>Creatinine 53 umol/L<br>PT; NT | Haemoglobin 132 g/l<br>lymphocyte count 1<br>neutrophil count 9.1<br>platelet count 203<br>**CRP 47 mg/L**<br>D-dimer; NR<br>Ferritin; NR<br>Creatinine 67 umol/L<br>PT; NR | Haemoglobin 147 g/l<br>**lymphocyte count 0.46**<br>neutrophil count 6.4<br>platelet count 154<br>**CRP 168 mg/L**<br>**D-dimer 3748 ug/l**<br>Ferritin; NR<br>Creatinine 69 umol/L<br>PT 10.4 seconds | Haemoglobin 132 g/l<br>**lymphocyte count 0.76**<br>neutrophil count 3.67<br>platelet count 153<br>**CRP 62 mg/L**<br>**D-dimer 5422 ug/l**<br>Ferritin 381 ug/l<br>Creatinine 64 umol/L<br>PT 11.2 seconds |
| Brain Imaging | MRI; Multiple patchy, asymmetric periventricular and subcortical white matter lesions in bilateral cerebral hemispheres, midbrain, dorsal pons, right middle cerebellar peduncle, medulla and right cerebellar hemisphere. Mixed diffusivity exhibited. Radiological progression on subsequent imaging during hospital admission | MRI; Lesions in the limbic system, predominantly in the left amygdala and hippocampus with partial restricted diffusion. | MRI; White matter lesions in the left anterior limbic structures with foci of increased diffusivity suggesting cellular inflammation. | MRI; Restricted diffusion in the inferior medulla with enhancement. Small area of restricted diffusion in left middle cerebellar peduncle. MRI spine; Extensive spinal cord abnormality predominantly involving the cervical and lower thoracic regions including the conus in keeping with transverse myelitis | MRI; No acute abnormality. Marked parenchymal atrophy with medial bitemporal predominance. | CT: No acute abnormality. | MRI : Progression of inflammatory demyelinating plaques since 2015 |
| CSF examination | **White cell count 8** per μl or mm<sup>3</sup>, lymphocyte 8, red cells 1, **protein 45.1mg/dl**, glucose 4.1, Paired serum glucose 5.6 mmol/L. **Oligoclonal bands** present. Viral PCR for SARS-CoV-2/ enterovirus/HSV negative. NMDA-R / CASPR2 / LGI-1 / Glycine receptor antibodies all negative, paraneoplastic anti-neuronal antibodies negative, anti-MOG and AQP4 negative | White cell none, Red cells 9, protein; protein 340mg/dl, glucose 4.1, Paired serum glucose 5.4mmol/L. Oligoclonal bands negative. Viral PCR for SARS-CoV-2/ enterovirus/HSV negative. NMDA-R / CASPR2 / LGI-1 / Glycine receptor antibodies all negative, paraneoplastic anti-neuronal antibodies negative. | White cell none, Red cells 25, **protein 98.7mg/dl**, glucose 4.5, Paired serum glucose 5.8 mmol/L. **Oligoclonal bands** present. Viral PCR for SARS-CoV-2/ enterovirus/HSV negative. NMDA-R / CASPR2 / LGI-1 / Glycine receptor antibodies all negative, paraneoplastic anti-neuronal antibodies negative. | White cell 1, red cells 0, **protein 51.4mg/dl**, glucose 3.1mmol/L. **Oligoclonal bands** present (identical to serum). Viral PCR for SARS-CoV-2/ enterovirus/HSV negative. NMDA-R / CASPR2 / LGI-1 / Glycine receptor antibodies all negative, paraneoplastic anti-neuronal antibodies negative. | Not performed (known early-onset advanced Alzheimer's Dementia with further cognitive decline) | Not performed | Not performed |
| Neurological treatments; recovery | Corticosteroids; Intravenous Methylprednisolone | Benzodiazepine, Levetiracetam IV & maintenance | Intubation and sedation for status epilepticus, Empirical aciclovir, antibiotics until CSF results available. IV valproate & maintenance | Intravenous Methylprednisolone | Supportive | Supportive and antibiotics (due to advanced disorder, osteomyelitis and pressure sores) | Glatiramer acetate injections 3 times/weekly |
| Outcome status | Recovery with mild neurological deficits; mRS 1 | Partial recovery with mild neurological deficits; mRS 1 | Full clinical recovery | Partial recovery with neurological deficits; Moderate disability mRS 4 | Partial recovery with neurological deficits; Severe disability mRS 5 | Died | Partial recovery with moderate disability; mRS 3 |

Footnote: M = male, F = female, NR = no result, CRP = C-Reactive protein, mRS = modified Rankin Score, ESR = Erythrocyte sedimentation rate, PCR = polymerase chain reaction, NMDA-R = N-methyl-D-aspartate Receptor antibodies, LGI-1 = Leucine-rich glioma inactivated-1, CASPR2 = contactin-associate protein-like 2, MOG = myelin oligodendrocyte glycoprotein, AQP4 = Aquaporin-4, PT = Prothrombin time. Paraneoplastic anti-neuronal antibodies including GABA-B, γ-Aminobutyric acid-B receptor; AMPA, GluR1 and GluR2 subunits of the AMPA receptor. CT = computed tomography. MRI = magnetic resonance imaging, Lymphocyte, neutrophil and platelet count, numbers x 10 E9/L.

Three patients (aged range between 46 to 79 years, three females) demonstrated a range of inflammatory encephalopathies. Patient 21 was diagnosed with limbic encephalitis, patient 22 with inflammatory encephalopathy, patient 27 with acute demyelinating encephalomyelitis (ADEM). Patient 9 was diagnosed with non-inflammatory encephalopathy and Patient 5 with transverse myelitis. Whilst four patients (9, 21, 22 and 27) presented with acute onset delirium and altered mental status, patients 21 and 22 also suffered from status epilepticus, cognitive impairment (scored 19 and 20 on the Montreal Cognitive Assessment respectively) and amnesia. Patient 5 presented with quadriparesis and altered sensation at the cervical level. Oligoclonal bands and mildly raised proteins were detected in the cerebrospinal fluid (CSF) of Patient 5, 22 and 27, whilst intrathecal SARS-CoV-2, paraneoplastic and autoimmune encephalitis antibodies (including N-methyl-D-aspartate Receptor, Leucine-rich glioma inactivated-1, contactin-associate protein-like 2, γ-Aminobutyric acid-B receptor, GluR1 and GluR2 subunits of the AMPA receptor) were negative in all three patients. The MRI Head scans showed partial diffusion restriction in the limbic system of patient 21 (Figure 2) and persistent diffusion-weighted hyperintensities without overt restriction in patient 22, suggestive of cellular inflammation. Patient 27 had progressive patchy, asymmetric periventricular and subcortical white matter hyperintense foci with diffusion restriction in the cerebral hemispheres, right cerebellar hemisphere and brainstem, features in keeping with ADEM (Figure 2). The MRI Spine in Patient 5 showed extensive spinal cord abnormality involving the cervical, lower thoracic and conal regions in keeping with transverse myelitis. Small regions of diffusion restriction were also identified in the medulla and middle cerebellar peduncle. Patient 22 required a brief critical care admission for 4 days, Patient 5 and 27 was given a course of intravenous (IV) followed by oral corticosteroids. All five patients recovered from their acute respiratory illness sufficiently for hospital discharge. However, only Patient 22 made a full neurological recovery, whilst the remaining patients were identified as having persistent neurological disability at discharge.

**Figure 2:**
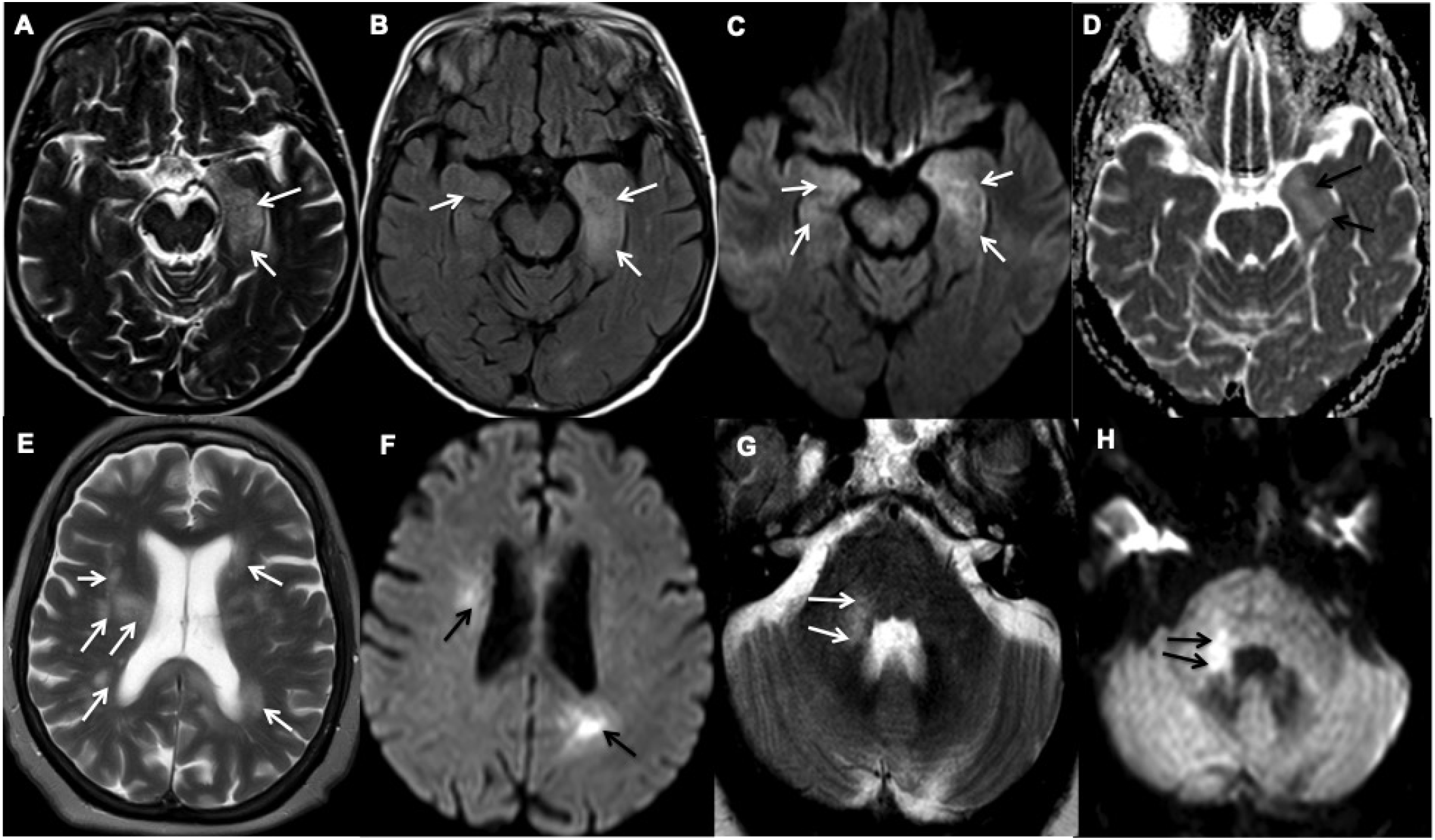
Representative examples of inflammatory encephalopathy. (A-D) Axial MR images of Patient 21 demonstrate limbic encephalitis. (A) T2-weighted and (B) Fluid attenuated inversion recovery (FLAIR) imaging demonstrate hyperintensity in both medial temporal lobes, but predominantly in the left amygdala and hippocampus (white arrows), (C) Diffusion-weighted imaging (white arrows) and (D) Apparent diffusion coefficient (ADC) show corresponding partial diffusion restriction (black arrows). (E-H) Axial MR images of Patient 27 demonstrate acute demyelinating encephalomyelitis. (E) T2-weighted imaging demonstrates patchy and asymmetric white matter hyperintensities within the periventricular and subcortical regions and (G) right middle cerebellar peduncle (white arrows). (F) and (H) Diffusion-weighted imaging shows corresponding high diffusion signal (black arrows).

#### Demyelination

(Patient 8, 14; Table 2)

Two male patients, (Patients 14 and 8), with known advanced multiple sclerosis (MS), (aged in their 50s and 60s respectively), presented with symptoms consistent with, and were diagnosed with COVID-19. Patient 8 presented with acute delirium whilst Patient 14 was hospitalized due to the MS relapse associated with worsening of his baseline MS-related limb weakness and dysarthria during the admission. The MRI Head performed showed progression of the inflammatory plaques since 2015 in Patient 14 but no acute changes were identified on the CT Head of Patient 8. None of them were admitted to the intensive care unit. Only Patient 14 survived with ongoing disability, whilst Patient 8 succumbed to septicaemia.

##### Movement Disorders

Myoclonus ± Opsoclonus (Patient 7, 19, 20; Table 3) Movement Disorders were seen in three patients with COVID-19 (Patients 7, 19 and 20; aged range between 57 to 86 years, two males). Patients 7 suffered from mild rest myoclonus of the arm, Patient 20 suffered from generalized rest myoclonus and Patient 19 was diagnosed with opsoclonus-myoclonus syndrome during the admission. No other neurological symptoms were identified. The CT/MRI Head demonstrated no acute abnormality in all patients. There was elevated CSF protein in Patient 19 and 20, with nothing else of note in the CSF analysis (including no detectable intrathecal SARS-CoV-2) in both patients. Normal electroencephalography (EEG) readings were demonstrated in Patient 20. All patients were treated with benzodiazepines and two patients made a full recovery at the time of discharge from hospital while Patient 19 had residual symptoms. Only Patient 7 required critical care admission for pulmonary COVID-19 symptoms.

**Table 3.**
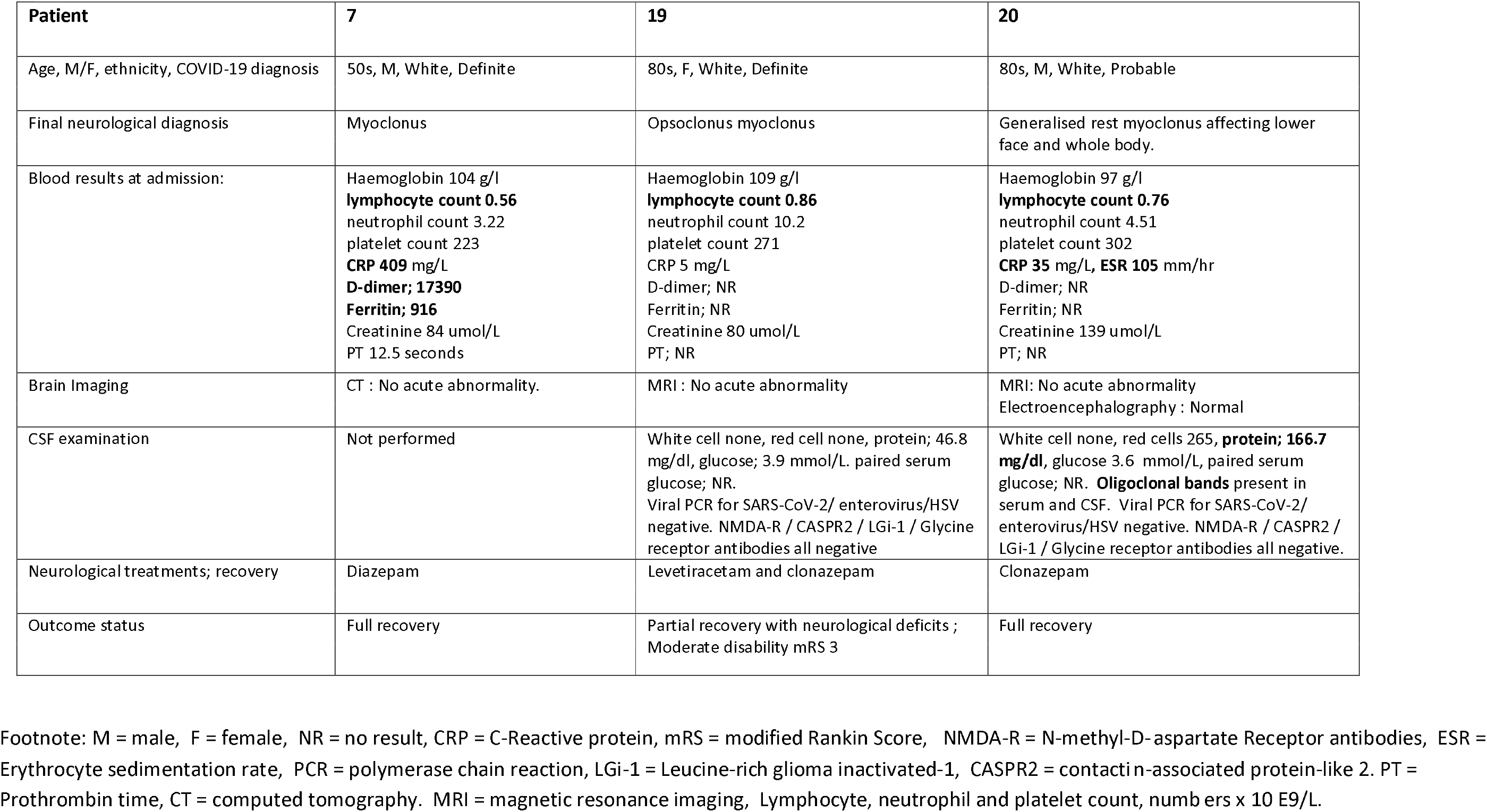
Three patients with movement disorders.

##### Peripheral Nervous System Disorders

2 acute inflammatory demyelinating polyneuropathy (Patient 13, 23) and 1 Brachial Plexopathy (Patient 26); (Table 4)

**Table 4.**
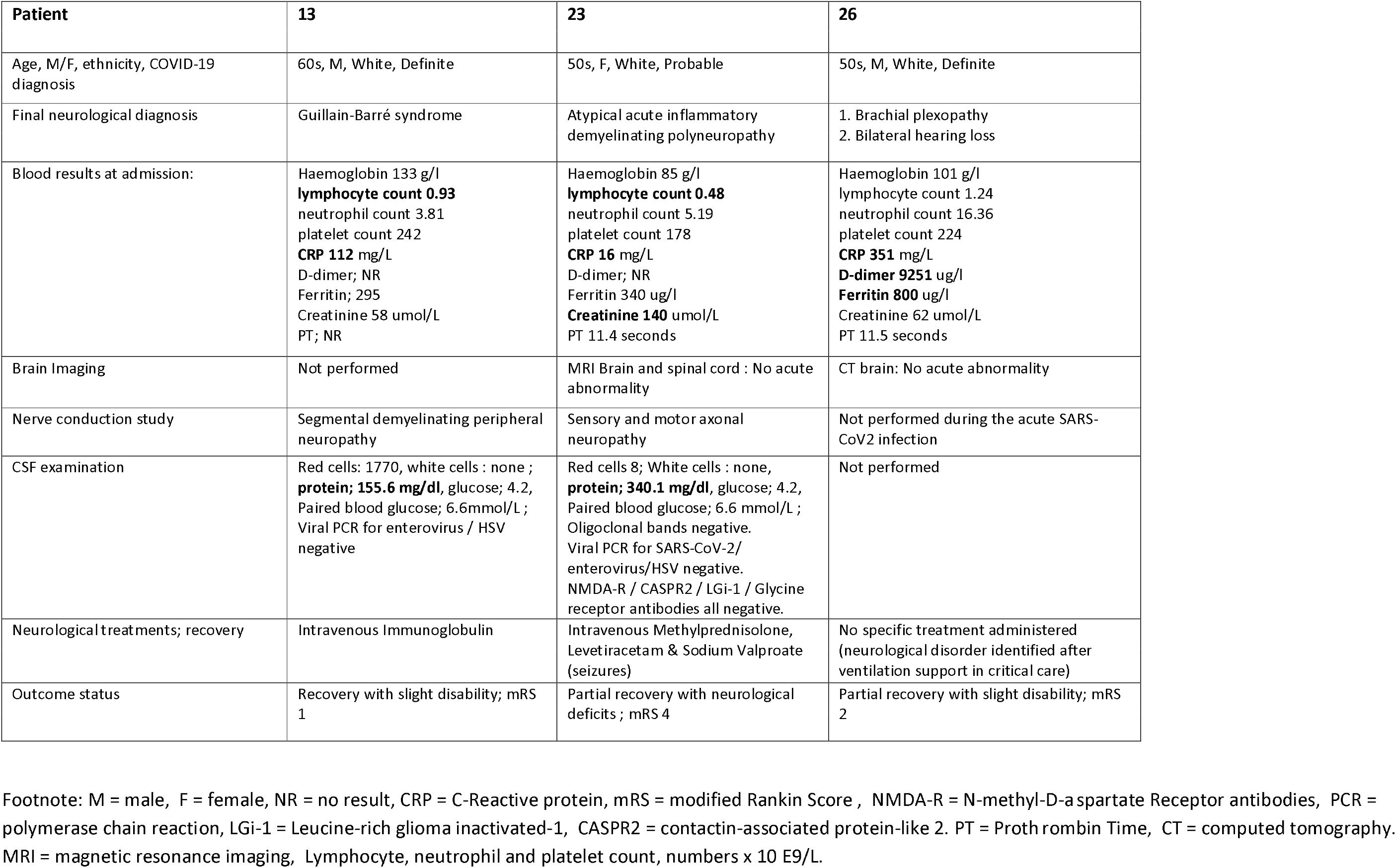
Three patients with disorders of peripheral nervous system.

Two COVID-19 patients (Patient 13 and 23; aged in their 50s and 60s respectively, two males) presented with ascending peripheral weakness and diagnosed with acute inflammatory demyelinating polyneuropathy or Guillain-Barré syndrome. The ascending distal limb weakness in patient 23 was associated with seizures and an atypical acute inflammatory demyelinating polyneuropathy was diagnosed. The nerve conduction studies confirmed features in keeping with segmental demyelinating peripheral neuropathy in Patient 13 and axonal neuropathy (motor and sensory) in Patient 23. Elevated CSF protein with otherwise normal CSF constituents was identified in both these patients. Patient 26 who presented with left upper limb weakness was diagnosed with brachial plexopathy.

Patient 13 was treated with IV immunoglobulin. Patients 23 and 26 required a short period of critical care admission and all patients have made a partial recovery. Patient 26 reported bilateral hearing loss following hospital discharge and has been referred for further investigation.

## Discussion

We report a variety of neurological disorders with a clinical impact in patients with COVID-19 infection admitted in a large tertiary institution during the ‘first wave’ of the COVID-19 pandemic in the UK. There have been isolated reports of various neurological disorders associated with previous outbreaks of the severe acute respiratory syndrome (SARS) and Middle East acute respiratory syndrome (MERS) (19, 20). Similarly, our cohort of cases demonstrates a wide range of COVID-19 related neurological disorders, ischaemic and haemorrhagic cerebrovascular events, inflammatory and non-inflammatory encephalopathy syndromes, transverse myelitis, movement disorders, and acute inflammatory demyelinating polyneuropathy with additional neurological manifestations. Excluding anosmia, the cumulative incidence of disorders is in 2.3% of our hospitalised patients with COVID-19, with 27% of them requiring critical care admission. The diagnosis of each of the neurological disorders was made in conjunction with a positive diagnosis of COVID-19, suggesting their association may not be fortuitous. Whilst recent case reports/series have described a range of neurological symptoms during the ongoing COVID-19 pandemic, there is often a lack of detailed clinical, radiological and laboratory findings of the neurological disorders amongst hospitalised patients with COVID-19, which reflect the challenge of studying the natural history of COVID-19 complications in this patient cohort.

Evidence from our cohort and recent studies have included non-specific initial presentations such as altered mental status or delirium, features commonly seen in the critically unwell with sepsis and hypoxaemia, as well as being potential early signs of dementia. The neurological disorders have been reported in patients who present solely with neurological signs and symptoms as well as those with established systemic or pulmonary illness related to COVID-19. These neurological features may precede or occur days after the onset of pulmonary symptoms. Hence, the variable and non-specific nature of the presentation and onset of the illness creates a diagnostic and therapeutic dilemma. Furthermore, the occasional delay of presentation and hospital admission during the first peak of the pandemic due to patients’ fear, isolation or shielding, may have lead to an increase in severity of the infection and neurological disorders at the point of diagnosis. Interestingly, there was no increased incidence or severity of the COVID-19 infection or neurological disorders amongst the BAME groups in our cohort. However, this could be due to the relatively small number of BAME communities in our geographical region.

There has been a reported increase in the incidence and severity of cerebrovascular disease associated with COVID-19, particularly in a younger cohort (6, 21). Our cohort demonstrated a high percentage of patients with acute ischaemic stroke and up to 38% of these had a large vessel occlusion. Some of the acute ischaemic cerebrovascular events with multifocal infarcts may be cardioembolic in nature, due to associated cardiovascular risk factors, but coagulopathy, vasculitis and viral endothelialitis have also been reported as potential causes of multi-vessel stroke in patients with COVID-19 (21, 22). The hyper-inflammatory syndrome or ‘cytokine storm’ strongly associated with severe COVID-19 infection could also contribute to the underlying aetiology (13).

Thrombotic microangiopathy and endothelial dysfunction, also evident in multiple organ systems related to COVID-19, may be contributory factors in sepsis/critical illness-related cerebral microbleeds (22, 23). The SARS-CoV-2 has been shown to preferentially bind to the angiotensin converting enzyme (ACE)-2 receptors that can be found in the endothelial lining, leading to the breakdown of the blood-brain barrier (8). However, cerebral microbleeds have similarly been reported in acute respiratory distress syndrome patients with a resemblance seen in cerebral microbleeds-related high altitude exposure, sharing a common underlying aetiology of hypoxaemia (24). This could likewise explain the findings in our cases with haemorrhagic neurological manifestations in COVID-19. Interestingly, both patients with isolated intraventricular haemorrhage had normal coagulation parameters, and the observed cerebral microbleeds were atypical for hypertensive or amyloid angiopathy causes. Other variables that may influence the presence and/or extent of microhaemorrhage in patients with COVID-19 include therapeutic anticoagulation and raised cerebral venous pressure secondary to ventilator measures in optimising patient oxygenation in the critical care setting (18).

The neurotropic potential of COVID-19 via direct viral axonal injury has been alluded to following scarce reports of the SARS-CoV-2 being detected in the CSF of patients with meningo-encephalitis and in animal models (25, 26). Similarly, few case reports have demonstrated imaging features of direct neuronal injury of the olfactory pathway in COVID-19 patients presenting with anosmia, adding strength to this potential mechanism (27). Nonetheless, no detectable intrathecal coronavirus strain was identified in our patients who presented with encephalopathy.

Interestingly, two patients who required critical care admission reported new onset hearing loss following hospital discharge, despite no acute abnormality reported on the admission MRI Head. A recent case report also described a possible association between sensorineural hearing impairment and COVID-19 in the critical care setting (28). It is postulated that such an observation could be due to the underlying hyperinflammatory process and/or the neuro-invasive potential of the SARS-CoV-2 against the auditory nervous system. Hence, it will be important to consider screening patients with severe COVID-19 infection for hearing impairment during the hospital admission.

An immunological response secondary to the SARS-CoV-2, resulting in cerebral inflammation and oedema with clinical encephalopathic features may offer an alternative explanation for the incidence of inflammatory and auto-immune encephalopathy disorders (14, 29). Antibodies against neuronal synaptic proteins have been demonstrated in autoimmune encephalitis, and there have been increased numbers of antibodies reported against other coronavirus strains, suggesting a possible association between auto-immune or inflammatory encephalopathic disorders and the COVID-19 infection (30, 31). Furthermore, the presence of both intrathecal and serum oligoclonal bands in two patients with acute encephalopathy and a patient with ADEM suggests that the immune-mediated response is not restricted to the intrathecal production of immunoglobulins. Post-infectious autoimmune disorder is also demonstrated in our case cohort of acute inflammatory demyelinating polyneuropathy, whereby the onset of neurological symptoms followed an initial period of illness related to COVID-19. Expected electrophysiological changes in keeping with demyelinating peripheral neuropathy was confirmed in one patient and the anticipated response to the IV immunoglobulin therapy was observed.

Limitations of our study include its lack of pathological evidence to prove causality. Furthermore, we only included hospitalised patients with COVID-19 in our study, thereby potentially underestimating the true incidence of the neurological associations in patients in the community. There were also inherent drawbacks in the sensitivity and specificity of the available RT-PCR swab tests during the study period, which may have underestimated the incidence of COVID-19 in the patient population (32).

Although the exact mechanism and possible causality of the SARS-CoV-2 infection associated with each of the presented neurological disorders remains unclear, it is likely that shared pathophysiological mechanisms are responsible for the various neurological manifestations of COVID-19. Our study lends further support to the growing body of evidence, aiding better understanding of the neurological features and optimizing management strategies using an approach guided by the evolution of clinical, laboratory and imaging features. Longitudinal follow-up of these patients is required to determine the long term effects, treatment response and outcome of the SARS-CoV-2 infection.

## Data Availability

Data is available upon request.

## Notes

Other disclosures or conflict of interest : None

### Competing Interest Statement

The authors have declared no competing interest.

### Funding Statement

AAH is partly funded by the Medical Research Council UK. No external funding declared.

### Author Declarations

This study was registered with and approved by the East Midlands-Derby Research Ethics Committee (Ref:18/EM/0292).

